# Detection of anti-premembrane antibody as a specific marker of four flavivirus serocomplexes and its application to serosurveillance in endemic regions

**DOI:** 10.1101/2023.09.21.23295701

**Authors:** Guan-Hua Chen, Yu-Ching Dai, Szu-Chia Hsieh, Jih-Jin Tsai, Ava Kristy Sy, Mario Jiz, Celia Pedroso, Carlos Brites, Eduardo Martins Netto, Phyllis J. Kanki, Danielle R. D. Saunders, Dana L. Vanlandingham, Stephen Higgs, Yan-Jang S. Huang, Wei-Kung Wang

## Abstract

In the past few decades, several emerging/re-emerging mosquito-borne flaviviruses have resulted in disease outbreaks of public health concern in the tropics and subtropics. Due to cross-reactivities of antibodies recognizing the envelope protein of different flaviviruses, serosurveillance remains a challenge. Previously we reported that anti-premembrane (prM) antibody can discriminate between three flavivirus infections by Western blot analysis. In this study, we aimed to develop a serological assay that can discriminate infection or exposure with flaviviruses from four serocomplexes, including dengue (DENV), Zika (ZIKV), West Nile (WNV) and yellow fever (YFV) viruses, and explore its application for serosurveillance in flavivirus-endemic countries. We employed Western blot analysis including antigens of six flaviviruses (DENV1, 2 and 4, WNV, ZIKV and YFV) from four serocomplexes. We tested serum samples from YF-17D vaccinees, and from DENV, ZIKV and WNV panels that had been confirmed by RT-PCR or by neutralization assays. The overall sensitivity/specificity of anti-prM antibodies for DENV, ZIKV, WNV, and YFV infections/exposure were 91.7%/96.4%, 91.7%/99.2%, 88.9%/98.3%, and 91.3%/92.5%, respectively. When testing 48 samples from Brazil, we identified multiple flavivirus infections/exposure including DENV and ZIKV, DENV and YFV, and DENV, ZIKV and YFV. When testing 50 samples from the Philippines, we detected DENV, ZIKV, and DENV and ZIKV infections with a ZIKV seroprevalence rate of 10%, which was consistent with reports of low-level circulation of ZIKV in Asia. Together, these findings suggest that anti-prM antibody is a flavivirus serocomplex-specific marker and can be employed to delineate four flavivirus infections/exposure in regions where multiple flaviviruses co-circulate.

## Introduction

In the genus *Flavivirus* of the family *Flaviviridae*, there are several mosquito-borne viruses causing significant diseases in humans; including the four serotypes of dengue virus (DENV) in the DENV serocomplex, West Nile virus (WNV) and Japanese encephalitis virus (JEV) in the JEV serocomplex, Zika virus (ZIKV), and yellow fever virus (YFV) as a single member [1].

The four serotypes of DENV (DENV1-DENV4) continue to be a global public health threat in tropical and subtropical regions [2-4]. It has been estimated that approximately 390 million DENV infections occur annually worldwide [2-4]. Most DENV infections are inapparent or subclinical with about 25% leading to clinical disease including dengue, dengue with warning signs, and severe dengue [2-4]. Of the DENV vaccine candidates that have completed different phases of clinical trials, Dengvaxia, a chimeric yellow fever-dengue tetravalent vaccine, was the first DENV vaccine licensed [5]. As DENV-seronegative children receiving Dengvaxia were reported to have a higher risk for hospitalization and severe dengue during subsequent DENV infection, Dengvaxia was recommended for DENV-seropositive individuals aged 9–45 years [5-7]. Pre-vaccination screening strategies using assays with high sensitivity and specificity have been proposed, highlighting the need for reliable serological tests to determine DENV serostatus in flavivirus-endemic regions [8].

Three additional flaviviruses, YFV, WNV, and ZIKV were also included in this research. In sub-Sahara Africa and tropical America, YFV is endemic with an estimate of 200,000 severe cases and up to 60,000 deaths per year [9,10]. The recent outbreaks in Angola and the Democratic Republic of the Congo, followed by the outbreaks in Brazil and Nigeria, suggest that YFV has expanded to new areas to affect large populations in South America and Africa [9,10]. First isolated in Uganda in 1937, WNV caused human cases and outbreaks in Africa and Europe; since 1999 WNV has spread throughout the continental U.S. and to Canada and Mexico [1,11]. The reports of increased incidence in the geographic distribution of WNV and travel-related WNV cases posed new challenge of serosurveillance for flaviviruses [11,12]. ZIKV, first isolated in Uganda in 1947, was associated with relatively few human cases until the outbreaks on Yap Island in 2007 and French Polynesia in 2013−2014. The subsequent explosive spread in the Americas since 2015 has resulted in ∼800,000 suspected or confirmed cases [13,14]. The association of ZIKV with microcephaly and other birth defects, known as congenital Zika syndrome (CZS), has raised global public health concern [13-15]. Despite a decline in ZIKV transmission since late 2017, the specter of its re-emergence and CZS in the endemic regions remains.

Knowing the seroprevalence rates of flaviviruses is critical to our understanding of the epidemiology and transmission dynamics of flaviviruses and critical for the development of intervention strategies. In addition, information about DENV serostatus can be used to evaluate DENV vaccine efficacy and determine if an individual would benefit from Dengvaxia and/or other vaccine candidates. Due to the presence of mosquito vectors in the regions, there is considerable geographic overlap in the distribution of different flaviviruses, such as DENV, JEV and ZIKV in Southeast Asia; DENV, YFV and ZIKV in South America; and DENV, YFV, WNV and ZIKV in sub-Sahara Africa. Our knowledge about the effects of prior immunity to one flavivirus on disease outcome of infection with another flavivirus in humans was primarily based on cohort studies. It has been reported that preexisting JEV neutralizing antibodies increased symptomatic DENV infection in Thailand [16]. Since the ZIKV outbreak during 2015−2017, two studies reported that prior DENV infection was associated with reduced risk of symptomatic ZIKV infection [17,18]. Another study showed that one prior ZIKV infection or one prior DENV followed by one ZIKV infection increased the risk of subsequent symptomatic DENV2 infection and severe disease, whereas a prior ZIKV with two or more DENV infections had a protective effect [19], underscoring the importance of reliable serological tests that can discriminate DENV, ZIKV and/or other flavivirus infections to improve our understanding of the complex interactions between DENV, ZIKV and/or other flaviviruses in endemic regions.

Present on the surface of flavivirus particles, the envelop (E) protein is the major target of neutralizing antibodies and vaccine development [1,20]. The ectodomain of the E protein contains 3 domains; the fusion loop (FL) is located at the tip of domain II and contains several highly conserved residues [1,20]. The premembrane (prM) protein, a glycoprotein of ∼19 kDa, is cleaved by furin or furin-like protease to precursor (pr) and membrane (M) proteins during maturation in the *trans*-Golgi [1,20]. Several serological tests have been developed based on the E protein including recombinant E protein, inactivated virions, or virus-like particles (VLPs) [20-23]. Due to cross-reactivity of anti-E antibodies to different flaviviruses, E protein-based serological tests cannot discriminate different flavivirus infections [20,23-27]. Nonstructural protein 1 (NS1)-based serological tests including enzyme-linked immunosorbent assay (ELISA), blockade of binding ELISA, and microsphere immunoassay have shown improved specificity [28-32]. However, the reduced durability of anti-NS1 antibodies could be a challenge for seroprevalence studies.

The plaque reduction neutralization test (PRNT) is considered the gold standard serological test and has been employed to confirm different flavivirus infections in serosurveillance and seroprevalence studies [1,23]. For individuals experiencing a single flavivirus infection, it identifies monotypic neutralizing antibodies against a single flavivirus, such as neutralizing antibody against one DENV serotype or ZIKV in individuals with primary DENV (pDENV) or primary ZIKV (pZIKV) infection, respectively [23,33-35]. For individuals experiencing multiple flavivirus infections, it reveals multitypic neutralizing antibodies against multiple flaviviruses, such as two or more DENV serotypes and/or other flaviviruses in individuals with secondary DENV (sDENV) infection [23,33-35]. Therefore, multitypic neutralizing antibodies were interpreted as unspecified flavivirus infections and cannot discriminate between the flaviviruses experienced in the past such as sDENV infection versus previous DENV and ZIKV (DENV+ZIKV) infections [23].

Previously, we employed Western blot analysis using an antigen panel of six flavivirus-infected cell lysates (DENV1-4, WNV and ZIKV) from three serocomplexes to test different panels with known flavivirus infections and reported that anti-prM antibodies can discriminate between DENV, ZIKV and WNV infections [36]. Whether or not this assay can be extended to include other flaviviruses such as YFV and applied to serosurveillance in flavivirus-endemic regions remains unanswered. In this study, we aimed to develop a serological test that can discriminate infection of flaviviruses from four serocomplexes including DENV, ZIKV, WNV and YFV, and examine samples collected from serosurveillance in the Philippines and Brazil. The underlying hypothesis was that detection of anti-prM antibody can discriminate infections caused by four flavivirus serocomplexes. We found anti-prM antibody is a specific marker for four flavivirus serocomplexes.

## Materials and methods

### Human samples

This study was approved by the Institutional Review Boards (IRBs) of the University of Hawaii (CHS#17568, 2022-00201, 2021-00947), the Kaohsiung Medical University Hospital, Taiwan (KMUH-IRB-960195, KMUH-IRB-E[I]-20170185), and the Research Institute for Tropical Medicine (RITM), Philippines (2019-042). The numbers, sampling time, sources and confirmation methods of different panels of serum or plasma samples with known flavivirus infections or vaccination are summarized in Table S1. Samples from reverse transcription-polymerase chain reaction (RT-PCR)-confirmed Zika cases including previously DENV-naïve (n=18) or DENV-exposed (n=13), designated as pZIKV or DENV+ZIKV panels respectively, were collected between July and March 2017 from the Pediatric Dengue Cohort Study and the Pediatric Dengue Hospital-based Study in Managua, Nicaragua [37,38]. The studies were approved by the IRBs of the University of California, Berkeley, and the Nicaraguan Ministry of Health. Samples from a ZIKV study in Salvador, an epicenter of ZIKV outbreak in Brazil, were confirmed by microneutralization tests (to ZIKV and DENV) as pZIKV (n=5), DENV+ZIKV (n=12), pDENV (n=4), and sDENV (n=21) panels as described previously [39]. The study was approved by Approved the Comitê de Ética em Pesquisa da Maternidade Climério de Oliveira/UFBA, Brazil (CAAE: 25336819.3.0000.5543/4.691.233, 2019). Samples (n=18) from blood donors, who tested positive for WNV transcription-mediated amplification (TMA), IgM and IgG antibodies, were designated as the WNV infection panel, were provided by the American Red Cross at Gaithersburg, Maryland [40]. Samples from a DENV seroprevalence study in Kaohsiung, Taiwan were confirmed by a microneutralization test (to DENV) as pDENV (n=17), sDENV (n=29) or DENV-naive (n=29) [41,42]; the sampling time was available based on questionnaires from study participants. Samples of YF-17D vaccinees were from the US (n=10) and Brazil (n=9) based on history of YF-17D vaccination [43]; samples from non-human primates (NHP) receiving YF-17D vaccine (n=4) were from the BEI Resources (NIAID, NIH). Additionally, samples from 50 participants (aged 2 to 56 years) collected between January 2018 and May 2019 from a fever surveillance program at the RITM, Philippines were included as a test panel [43]. These were non-Dengvaxia recipients and presented with symptoms suspected of dengue; blood samples were negative by DENV RT-PCR or DENV IgM-capture ELISA. Another panel of samples (n=48) of suspected ZIKV cases (aged 15−70 years) collected between 2015 and 2016 from the ZIKV study in Salvador, Brazil was included as a second test panel [31].

### Western blot analysis

Uninfected (mock) Vero cells and Vero cells infected with DENV1 (Hawaii strain), DENV2 (NGC strain), DENV4 (H241 strain), ZIKV (PRVABC59 strain), WNV (NY99 strain) or YFV (17D vaccine strain) were lysed with NP-40 lysis buffer (1% NP-40, 50 mM Tris pH 8.0, 150 mM NaCl, 2 mM EDTA, and 1 mM Na_3_VO_4_) when 50% of cells were found to have cytopathic effects. The cell lysates were loaded into two half-gels (seven wells each) and subjected to SDS-12% polyacrylamide gel electrophoresis under non-reducing condition (2% SDS, 0.5 M Tris pH 6.8, 20 % glycerol, 0.001 % bromophenol blue, final) [24,36], followed by transfer to nitrocellulose membrane (Trans-Blot Turbo RTA Midi Transfer Kit, BioRad), hybridization with human serum/plasma samples (1:200 dilution) or mouse monoclonal antibody (mAb) and secondary antibody (IRDye® 800CW-conjugated goat anti-human IgG at 1:10000). The signals were detected by Li Cor Odyssey classic (LiCor Biosciences) and analyzed by Image Studio software with both short and long exposures [36,42]. Each gel was read independently by two researchers with the results summarized in Supplementary Table 2. To test the stability of the assay, the half-membranes after blocking step were stored in the -20°C freezer until use for hybridization to serum/plasma.

### Expression of YF-17D prM/E proteins

293T cells (1x10^5^ cells) were transfected with 10 μg of a plasmid expressing the prM/E proteins of YF-17D. At 48 h, cells were washed with 1X PBS and treated with 1% NP40 lysis buffer, followed by centrifugation at 20,000 × g at 4°C f or 30 min to obtain cell lysates for Western blot analysis probed with human dengue-immune serum as described above [24,36].

### DENV FL-VLP IgG ELISA

IgG ELISA using DENV1 FL-mutated VLPs was described previously [44]. Briefly, DENV1 FL-mutated VLPs (containing W101A and F108A mutations) were coated onto 96-well plates at 4°C overnight, followed by blocking (StartingBlock blocking buffer, Thermo Scientific, Waltham, MA) at room temperature for 1 h, incubation with primary antibody (serum or plasma at 1:400 dilution) at 37°C for 2 h, wash with washing buffer (0.5% Tween-20 in 1X PBS) 4 times, incubation with secondary antibody (anti-human IgG conjugated with horseradish peroxidase [HRP] at 1:10,000 dilution, Jackson Immune Research Laboratory, West Grove, PA) at 37°C for 1 h, and wash with washing buffer 6 times [40,44]. After incubation with tetramethylbenzidine substrate (Thermo Scientific, Waltham, MA) at room temperature for 15 min and stop solution, the OD at 450 nm was read with a reference wavelength of 630 nm. Each ELISA plate contained two positive controls (OD higher than 1; two confirmed-DENV samples), four negative controls (DENV-naïve sera or plasma), and test samples (all in duplicate). For comparison between plates, the relative OD (rOD) values were calculated by the OD values divided by the mean OD value of one positive control (OD close to 1) in the same plate. The cutoff rOD was defined by the mean rOD value of negatives plus 12 standard deviations, which gave a confidence level of 99.9% from 4 negatives [45]. Each ELISA was performed in duplicate.

### Microneutralization test

Microneutralization tests were performed as described previously [39,42]. Briefly, two-fold serial dilutions of serum were mixed with 50 focus-forming units of DENV1 (Hawaii), DENV2 (NGC), DENV3 (CH53489), DENV4 (H241), ZIKV (PRVABC59), or YFV (YF-17D) at 37°C for 1 h; the mixtures were added to each well of 96-well plate which was pre-seeded with Vero cells (3 x 10^4^ cells per well) one day prior to infection. This was followed by incubation at 37°C for 48-70 h, removal of medium, fixation [39,42], mouse mAb 4G2 and secondary antibodies (IRDye® 800CW-conjugated goat anti-mouse IgG and DRAQ5™ fluorescent probe at 1:10000). The signals (800 nm/700 nm fluorescence) were detected using a LiCor Odyssey imager (LiCor Biosciences) and analyzed by Image Studio to determine percent neutralization at different concentrations and 90% neutralization (NT_90_) titers [39,42].

### Statistical analysis

The sensitivity, specificity and 95% confidence intervals (CI) were calculated by Excel. The two-tailed Fisher’s exact test and two-tailed Mann-Whitney test were used to compare categorical and quantitative variables, respectively, between two groups (GraphPad Prism 6). The positive, negative and overall agreements and kappa assessment were calculated by the SPSS 20.

## Results

### Antibody response following YF-17D vaccination

The antigen panel in Western blot analysis included lysates derived from Vero cells infected with six flaviviruses (DENV1, 2 and 4, WNV, ZIKV and YF-17D vaccine strain) from four serocomplexes. The control panels with known flavivirus infections or vaccination included serum or plasma samples from YF-17D vaccinees (YF-17D panel), RT-PCR-confirmed ZIKV cases (pZIKV and DENV+ZIKV panels), TMA-confirmed WNV infection (WNV panel), and neutralization-confirmed DENV infection or DENV-naïve participants (pDENV, sDENV and DENV-naïve panels) from a DENV seroprevalence study (Table S1). We first examined antibody response in two vaccinees ∼3 months following YF-17D vaccination by Western blot analysis. Each lane contained individual viral proteins presumably in equal molar ratio except for those structural proteins released with virions; a previously described flavivirus group-reactive mouse mAb FL0232, which recognized E proteins (DENV1-4, ZIKV, WNV and YFV) equally well, was used to verify comparable amounts of loaded antigens (Figure 1(C)) [23,36]. DENV3-infected cell lysate was not included due to high amino acid homology between DENV3 and DENV1 (compared with DENV2 or DENV4) and the convenience of loading seven lanes in a half membrane. As shown in Figure 1(A), anti-E antibodies cross-reactive to all six flaviviruses tested, and anti-NS1 and anti-prM antibodies recognizing YFV only were observed. A similar trend was observed in 17 other samples, although some recognized YFV E protein only (Table S2). Notably, the YFV prM protein migrated at a slower rate than the prM proteins of other flaviviruses (DENV1, 2 and 4, WNV and ZIKV) tested, corresponding to the size of a 23 kDa protein; this was confirmed by the size of prM protein expressed by a YF-17D prM/E plasmid (Figure 1(B)). We next examined samples from three NHPs receiving YF-17D vaccine (1 to 18 months, pooled sera); anti-E antibodies recognizing YFV and/or other five flaviviruses tested together with anti-NS1 and anti-prM antibodies recognizing YFV only were observed (Figure 1(D)). As a comparison, no protein band corresponding to E, NS1 or prM protein was recognized by a DENV-naive sample (Figure 1(E), Table S2). Different viral protein bands recognized by samples from 19 YF-17D vaccinees and four NHPs receiving YF-17D vaccine are summarized in Table 1.

**Figure 1.**
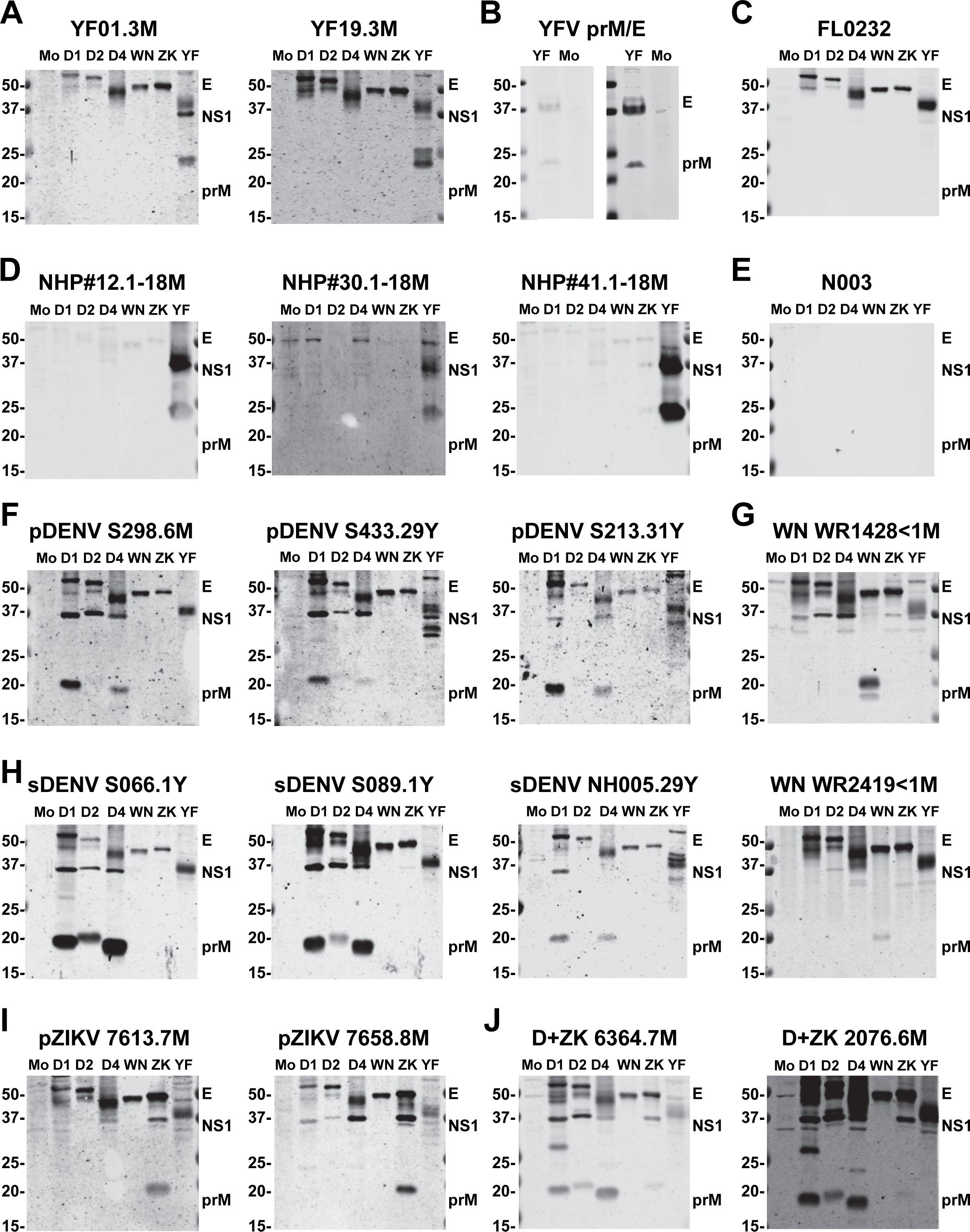
Antibody response to six flavivirus antigens following YF-17D vaccination or DENV, ZIKV or WNV infection. Lysates derived from mock-, DENV1-, DENV2-, DENV4-, WNV-, ZIKV-, and YFV (17D strain)-infected Vero cells were subjected to SDS-12% polyacrylamide gel electrophoresis under non-reducing condition and Western blot analysis probed with different serum/plasma samples or anti-E mouse mAb FL0232 (C). Results of YF-17D vaccinees (A) and NHPs receiving YF-17D vaccine (D), DENV-naïve participant (E), and participants with pDENV (F), WNV (G), sDENV (H), pZIKV (I), and DENV+ZIKV (D+ZK) (J) infections. Lysates derived from 293T cells transfected with YF-17D prM/E plasmid were subjected to Western blot analysis and probed with a DENV- and YFV-immune serum, short (left) and long (right) exposure (B). The sampling time post-symptom onset, vaccination or TMA test was indicated after sample ID. pDENV, primary DENV infection; sDENV, secondary DENV infection; pZIKV, primary ZIKV infection. The positions of E, NS1 and prM protein bands are indicated. The size of molecular weight markers is shown in kDa. Mo: mock, D1: DENV1, D2: DENV2, D4: DENV4, WN: WNV, ZK: ZIKV, and YF: YF-17D.

**Table 1.** Summary of viral proteins recognized by different panels^a^ in Western blot analysis.

| Protein bands recognized<sup>c,d</sup> | No. of positive/total samples (%) in different serum/plasma panels |  |  |  |  |  |  |
| --- | --- | --- | --- | --- | --- | --- | --- |
|  | DENV-naive | pDENV | sDENV | pZIKV | DENV+ZIKV | WNV<sup>b</sup> | YF-17D |
| D1, D2 or D4 NS1 | 6/29<br>(20.7%) | **16/21**<br>**(76.2%)** | **49/50**<br>**(98.0%)** | 20/23<br>(87.9%) | **25/25**<br>**(100%)** | 14/18<br>(77.8%) | 1/14<sup>g</sup><br>(7.1%) |
| ZIKV NS1 | 0/29<br>(0%) | 0/21<br>(0%) | 13/50<br>(26.0%) | **23/23**<br>**(100%)** | **25/25**<br>**(100%)** | 5/18<br>(27.8%) | 0/14<sup>g</sup><br>(0%) |
| WNV NS1 | 0/29<br>(0%) | 0/21<br>(0%) | 0/50<br>(0%) | 0/23<br>(0%) | 1/25<br>(4.0%) | **9/18**<br>**(50.0%)** | 0/23<br>(0%) |
| YFV NS1 | 7/29<br>(24.1%) | 5/17<sup>f</sup><br>(29.4%) | 6/29<sup>f</sup><br>(20.7%) | 3/18<sup>f</sup><br>(16.7%) | 10/13<sup>f</sup><br>(76.9%) | NA<sup>e</sup> | **22/23**<br>**(95.7%)** |
| D1, D2 or D4 prM | 0/29<br>(0%) | **17/21**<br>**(81.0%)** | **49/50**<br>**(98.0%)** | 2/23<br>(8.7%) | **22/25**<br>**(88.0%)** | 0/18<br>(0%) | 1/14<sup>g</sup><br>(7.1%) |
| ZIKV prM | 0/29<br>(0%) | 0/21<br>(0%) | 1/50<br>(2.0%) | **23/23**<br>**(100%)** | **21/25**<br>**(84.0%)** | 0/18<br>(0%) | 0/14<sup>g</sup><br>(0%) |
| WNV prM | 0/29<br>(0%) | 0/21<br>(0%) | 0/50<br>(0%) | 0/23<br>(0%) | 3/25<br>(12.0%) | **16/18**<br>**(88.9%)** | 0/23<br>(0%) |
| YFV prM | 1/29<br>(3.4%) | 1/17<sup>f</sup><br>(5.9%) | 2/29<sup>f</sup><br>(6.9%) | 2/18<sup>f</sup><br>(11.1%) | 2/13<sup>f</sup><br>(15.4%) | NA<sup>e</sup> | **21/23**<br>**(91.3%)** |
| any E (D1, D2, D4, ZIKV, WNV or YFV) | 0/29<br>(0%) | **21/21**<br>**(100%)** | **50/50**<br>**(100%)** | **23/23**<br>**(100%)** | **25/25**<br>**(100%)** | **18/18**<br>**(100%)** | **23/23**<br>**(100%)** |

<sup>a</sup>pDENV, primary DENV infection; sDENV, secondary DENV infection; pZIKV, primary ZIKV infection; DENV+ZIKV, previous DENV and ZIKV infections; WNV, WNV infection; YF-17D, YF-17D vaccination.
<sup>b</sup>Index samples tested positive for WNV transcription-mediated amplification, IgM and IgG from blood donors at the American Red Cross [40].
<sup>c</sup>NS1, nonstructural protein 1; prM, premembrane; D1, DENV1; D2, DENV2; D4, DENV4.
<sup>d</sup>No. (%) of homologous prM and NS1 proteins recognized by each panel are bolded.
<sup>e</sup>NA, not applicable; the history of YF-17D vaccination was not available in this panel.
<sup>f</sup>Due to the lack of history of YF-17D vaccination or infection from a subset of the pDENV (n=4), sDENV (n=21), pZIKV (n=5) and DENV+ZIKV (n=12) panels from Brazil, these samples were not included in the analysis of YFV NS1 or prM recognition.
<sup>g</sup>Due to the lack of history of DENV or ZIKV infection from a subset of the YF-17D (n=9) panel from Brazil, these samples were not included in the analysis of DENV and ZIKV NS1 or prM recognition.

### Anti-prM antibodies can discriminate four flavivirus infections or exposure

We further examined antibody response following DENV infection (Table S1). The results of three samples each from the pDENV panel and sDENV panel were shown in Figures 1(F) and 1(H), respectively. Anti-E antibodies cross-reactive to all six flaviviruses, anti-NS1 antibodies to one to three DENV serotypes with cross-reactivity to ZIKV or YFV, and anti-prM antibodies to DENV without cross-reactivity to ZIKV, WNV or YFV were found in both panels, including samples collected more than 29 years after infection (Figures 1(F) and 1(H)). A similar pattern of viral protein recognition was observed in other samples of the pDENV and sDENV panels (Table S2), except that the sDENV panel had a higher rate of cross-reactivity to ZIKV NS1 protein compared with the pDENV panel (26.0% vs. 0%, two-tailed Fisher’s exact test, *P*=0.007). As a comparison, WNV samples had anti-E antibodies cross-reactive to all six flaviviruses tested, anti-NS1 antibodies recognizing WNV and cross-reactive to DENV, ZIKV or YFV, and anti-prM antibodies recognizing WNV only (Figure 1(G), Table S2). We next examined two samples each from the pZIKV and DENV+ZIKV panels. Anti-E antibodies cross-reactive to all six flaviviruses tested and anti-NS1 antibodies to ZIKV with cross-reactivity to DENV (one to three serotypes) and YFV were observed in both panels. In contrast, anti-prM antibodies were found to recognize ZIKV only in the pZIKV panel and recognize both ZIKV and DENV in the DENV+ZIKV panel, as verified by long exposure (Figures 1(I) and 1(J), and data not shown). A similar trend was observed in other samples of the pZIKV and DENV+ZIKV panels (Table S2). Table 1 summarizes different viral protein bands recognized in 189 samples from the seven panels.

Table 2 summarizes the sensitivity and specificity of antibodies recognizing different NS1 or prM proteins to discriminate infection with DENV, ZIKV and WNV as well as vaccination with YF-17D. The overall sensitivity of anti-NS1 antibodies ranged from 50% to 100%, and the specificity 51.2% to 99.4%. In contrast, the overall sensitivity/specificity of anti-prM antibodies was higher (91.7/96.4%, 91.7/99.2%, 88.9/98.3%, and 91.3/92.5% for DENV, ZIKV, WNV, and YF-17D infection/vaccination, respectively), suggesting that anti-prM antibodies is a serocomplex-specific marker for the four flavivirus serocomplexes tested.

**Table 2.** Sensitivity and specificity of viral proteins recognized by different panels in Western blot analysis.

| Viral proteins recognized<sup>c</sup> | Group | % Sensitivity (95% CI)<sup>a,b</sup> | % Specificity (95% CI)<sup>a,b</sup> |
| --- | --- | --- | --- |
| D1, D2 or D4 NS1 | overall | 93.8 (88.9–96.2) | 51.2 (40.5–56.6) |
|  | subgroup | pDENV:76.2, sDENV:98.0, DENV+ZIKV:100 | DENV-naive:79.3, pZIKV:87.0, WNV:22.2, YF-17D:92.9 |
| ZIKV NS1 | overall | 100 (100–100) | 86.4 (80.5–89.4) |
|  | subgroup | pZIKV:100, DENV+ZIKV:100 | DENV-naive:100, pDENV:100, sDENV:74.0, WNV:72.2, YF-17D:100 |
| WNV NS1 | overall | 50.0 (26.9–61.8) | 99.4 (98.3–100) |
|  | subgroup | WNV:50.0 | DENV-naive:100, pDENV:100, sDENV:100, pZIKV:100, DENV+ZIKV:96.0, YF-17D:100 |
| YFV NS1 | overall | 95.7 (87.3–99.9) | 70.8 (62.1–75.2) |
|  | subgroup | YF17D:95.7 | DENV-naive:75.9, pDENV:70.6, sDENV:79.3, pZIKV:83.3, DENV+ZIKV:23.1, WNV:NA<sup>e</sup> |
| D1, D2 or D4 prM | overall | **91.7 (86.1–94.5)<sup>d</sup>** | **96.4 (92.5–98.5)<sup>d</sup>** |
|  | subgroup | pDENV:81.0, sDENV:98.0, DENV+ZIKV:88.0 | DENV-naive:100, pZIKV:91.3, WNV:100, YF-17D:92.9 |
| ZIKV prM | overall | **91.7 (83.9–95.7)<sup>d</sup>** | **99.2 (97.8–100)<sup>d</sup>** |
|  | subgroup | pZIKV:100, DENV+ZIKV:84.0 | DENV-naive:100, pDENV:100, sDENV:98.0, WNV:100, YF-17D:100 |
| WNV prM | overall | **88.9 (74.4–96.3)<sup>d</sup>** | **98.3 (96.3–99.3)<sup>d</sup>** |
|  | subgroup | pWNV:88.9 | DENV-naive:100, pDENV:100, sDENV:100, pZIKV:100, DENV+ZIKV:88.0, YF-17D:100 |
| YFV prM | overall | **91.3 (79.8–97.2)<sup>d</sup>** | **92.5 (87.4–95.0)<sup>d</sup>** |
|  | subgroup | YF17D:91.3 | DENV-naive:96.6, pDENV:94.1, sDENV:93.1, pZIKV:88.9, DENV+ZIKV:84.6, WNV:NA<sup>e</sup> |

<sup>a</sup>CI, confidence interval. pDENV, primary DENV infection; sDENV, secondary DENV infection; pZIKV, primary ZIKV infection; DENV+ZIKV, previous DENV and ZIKV infections; WNV, WNV infection; YF-17D, YF-17D vaccination.
<sup>b</sup>For simplicity, the 95% CIs in the subgroup are not shown.
<sup>c</sup>NS1, nonstructural protein 1; prM, premembrane; D1, DENV1; D2, DENV2; D4, DENV4.
<sup>d</sup>The sensitivity and specificity of recognizing prM protein are bolded.
<sup>e</sup>NA, not applicable; the history of YF-17D vaccination was not available in this panel.

### Serosurveillance by testing samples from endemic countries

To further examine whether or not this assay can be used for serosurveillance in countries where multiple flaviviruses are endemic and sympatric, we first tested 50 samples from a fever surveillance program in the Philippines between 2018 and 2019. Most samples had anti-E antibodies cross-reactive to all six flaviviruses, anti-NS1 antibodies to one to three DENV serotypes and/or ZIKV, and anti-prM antibodies to one to three DENV serotypes only, suggesting previous DENV infection (Figure 2(A)). Other samples did not have anti-E, anti-NS1 or anti-prM antibodies to DENV, ZIKV, WNV or YFV, suggesting that they were seronegative to these flaviviruses (Figure 2(E)). Three samples had anti-E antibodies cross-reactive to all six flaviviruses, anti-NS1 antibodies to one to three DENV serotypes and YFV or ZIKV, and anti-prM antibodies to one to three DENV serotypes and YFV without cross-reactivity to ZIKV or WNV, suggesting previous DENV and YFV infections/vaccination (Figure 2(B)). Four samples had anti-E antibodies cross-reactive to all six flaviviruses, anti-NS1 antibodies to one to three DENV serotypes and ZIKV or YFV, and anti-prM antibodies to one to three DENV serotypes and ZIKV without cross-reactivity to WNV or YFV, suggesting previous DENV and ZIKV infections (Figure 2(D)), whereas one sample had anti-E antibodies to ZIKV with faint cross-reactivity to DENV2, anti-NS1 antibodies to ZIKV with faint cross-reactivity to DENV4, and anti-prM antibodies to ZIKV only, suggesting pZIKV infection (Figure 2(C)).

**Figure 2.**
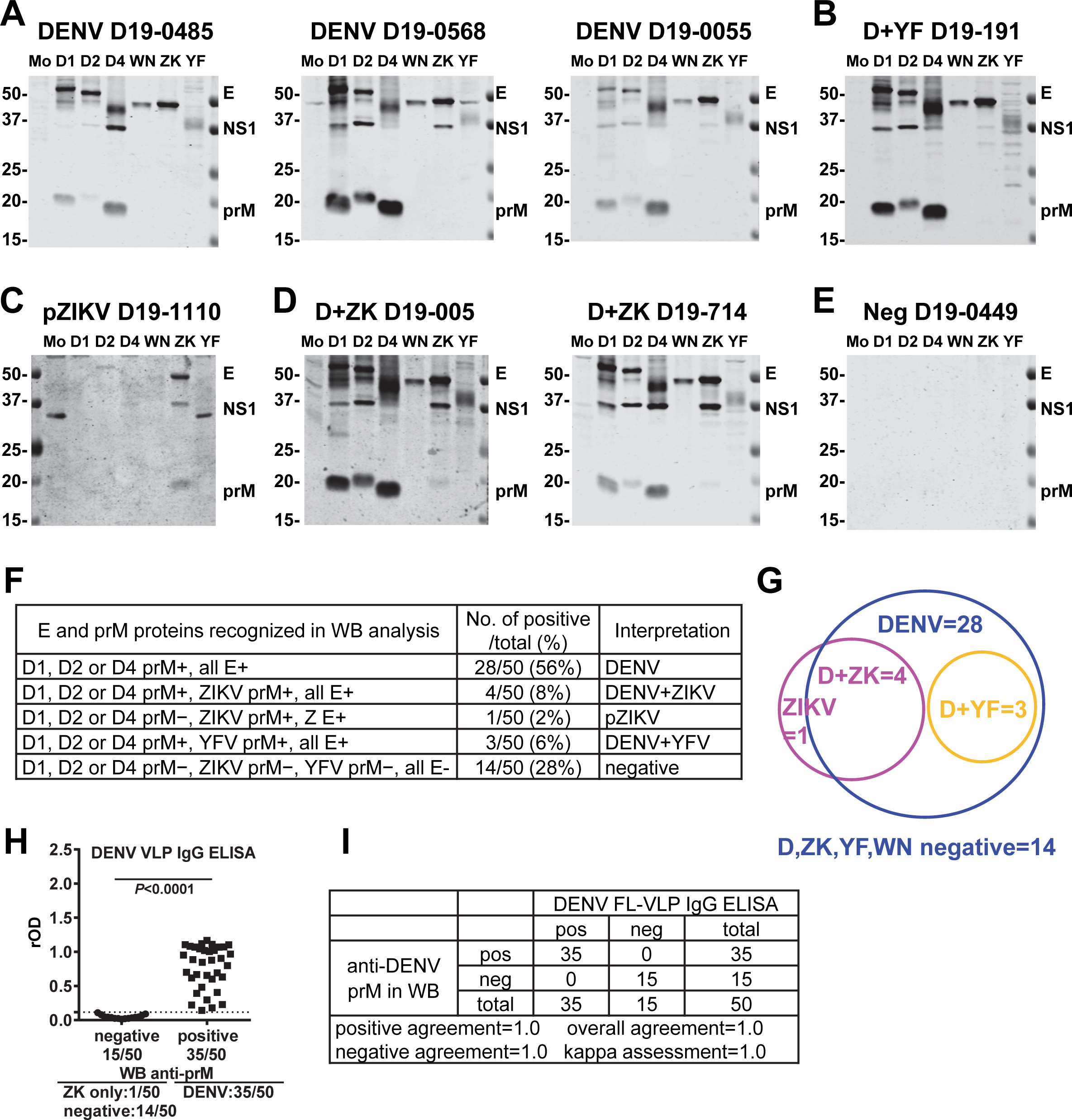
Antibody response to six flavivirus antigens in samples from a fever surveillance program in the Philippines. (A-E) Results of participants with previous DENV infection (A), previous DENV and YFV infections/vaccination (D+YF) (B), pZIKV infection (C), previous DENV and ZIKV infections (D+ZK) (D), and seronegative to DENV, ZIKV, YFV and WNV (Neg) (E). The positions of E, NS1 and prM protein bands are indicated. The size of molecular weight markers is shown in kDa. Mo: mock, D1: DENV1, D2: DENV2, D4: DENV4, WN: WNV, ZK: ZIKV, and YF: YF-17D. (F,G) The pattern of E and prM proteins recognized and the number/percentage of positive and total samples based on Western blot analysis (F) and a graphic summary (G). (H,I) Results of DENV FL-VLP IgG ELISA (H) and comparison with that of anti-DENV prM reactivity in Western blot analysis (I). rOD: the relative OD. The two-tailed Mann-Whitney test was performed in panel H.

The pattern of E and prM proteins recognized in Western blot analysis and the number/ percentage of positive and total samples from the Philippines are summarized in Figures 2(F) and 2(G). Of the 50 participants, 28 (56%) had previous DENV infection, 4 (8%) previous DENV and ZIKV infections, 1 (2%) previous ZIKV infection, 3 (6%) previous DENV and YFV infections/vaccination, and 14 (28%) negatives to the four flaviviruses tested; altogether 35 (70%) had previous DENV infection. We further tested with a previously reported IgG ELISA based on DENV FL-mutated VLP, and found 35 (70%) were positive and 15 (30%) negative (Figure 2(H)); this is consistent with the results of Western blot analysis. Comparing the Western blot analysis and DENV FL-VLP IgG ELISA, which had a sensitivity/specificity of 100.0%/93.3%, the positive, negative and overall agreements were all 1.0 with a kappa assessment of 1.0 (Figure 2(I)).

We next tested 48 samples from suspected Zika cases collected in Salvador, Brazil during the early phase of the ZIKV outbreak between 2015 and 2016. Anti-E antibodies cross-reactive to all six flaviviruses and anti-NS1 antibodies recognizing one to three DENV serotypes and/or ZIKV or YFV were found in all samples tested. Based on the recognition of anti-prM antibodies, we found four patterns. Some samples recognized DENV prM protein of one to three serotypes without cross-reactivity to ZIKV, YFV or WNV, suggesting previous DENV infection (Figure 3(A)), whereas others recognized DENV (one to three serotypes) and ZIKV or YFV prM proteins, suggesting previous DENV and ZIKV infections (Figure 3(B)) or DENV and YFV infections/vaccination (Figure 3(C)), respectively. Interestingly, some samples recognized DENV (one to three serotypes), ZIKV and YFV prM proteins, suggesting previous DENV, ZIKV and YFV infections/vaccination (Figure 3(D)).

**Figure 3.**
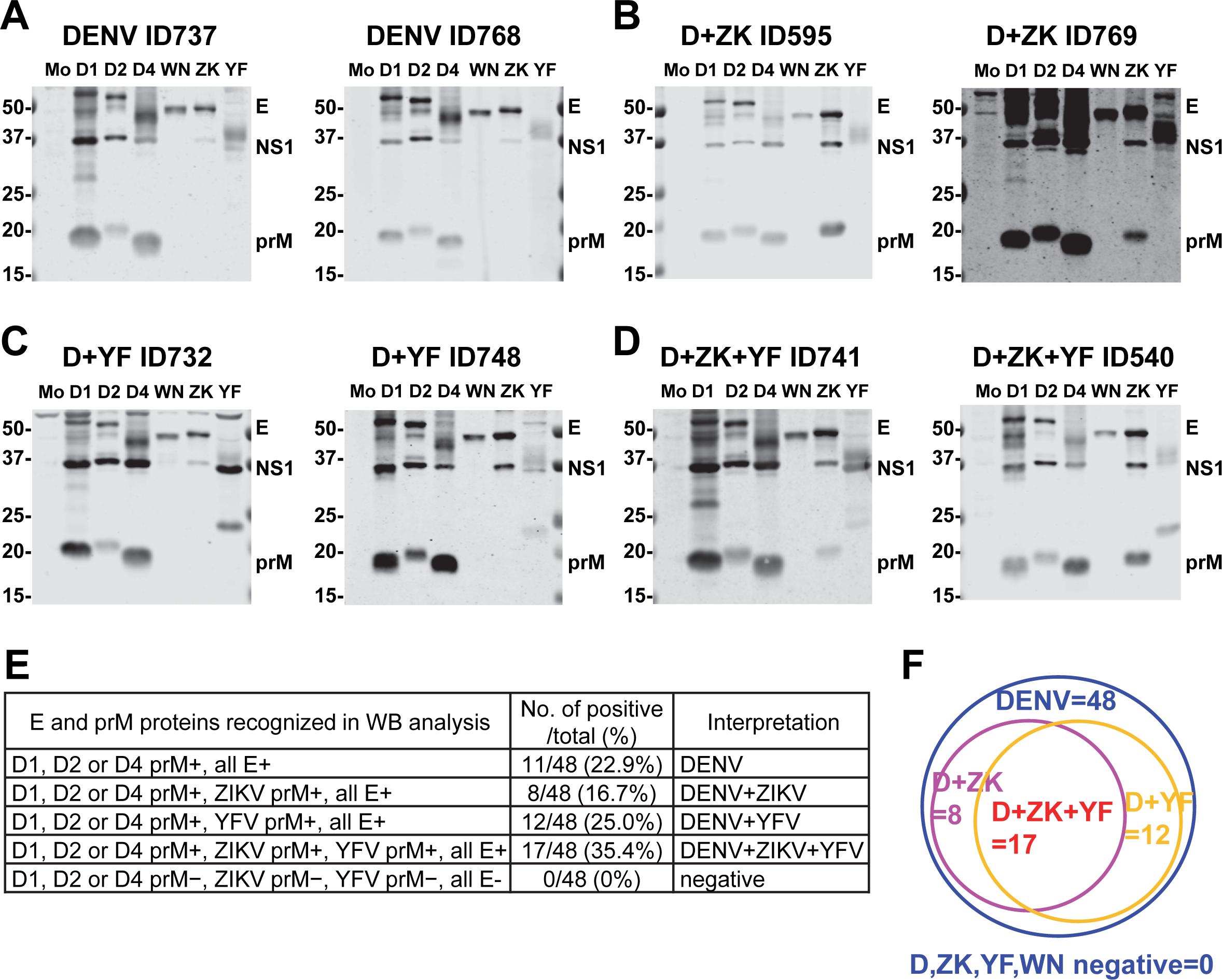
Antibody response to six flavivirus antigens in samples from suspected ZIKV cased during the ZIKV outbreak in Brazil. (A-D) Results of participants with previous DENV infection (A), previous DENV and ZIKV infections (D+ZK) (B) previous DENV and YFV infections/vaccination (D+YF) (C), and previous DENV, ZIKV and YFV infections/vaccination (D+ZK+YF) (D). The positions of E, NS1 and prM protein bands are indicated. The size of molecular weight markers is shown in kDa. Mo: mock, D1: DENV1, D2: DENV2, D4: DENV4, WN: WNV, ZK: ZIKV, and YF: YF-17D. (E,F) The pattern of E and prM proteins recognized and the number/percentage of positive and total samples based on Western blot analysis (E) and a graphic summary (F).

The pattern of E and prM proteins recognized in Western blot analysis and the number/percentage of positive and total samples from Brazil are summarized in Figures 3(E) and 3(F). Of the 48 participants, 11 (22.9%) had previous DENV infection, 8 (16.7%) previous DENV and ZIKV infections, 12 (25%) previous DENV and YFV infections/vaccination, and 17 (35.4%) previous DENV, ZIKV and YFV infections/vaccination. Taken together, all participants (100%) had previous DENV infection, 52.1% (25/48) previous ZIKV infection, and 60.4% (29/48) previous YFV infection or vaccination.

## Discussion

In this study, we employed antigens of six flaviviruses from four serocomplexes in Western blot analysis to test six panels of samples with well-documented flavivirus infections or vaccination and found that anti-prM antibodies is a specific marker for four flavivirus serocomplexes with an overall sensitivity/specificity of 91.7%/96.4%, 91.7%/99.2%, 88.9%/98.3%, and 91.3%/92.5%, for DENV, ZIKV, WNV, and YFV infections/vaccination, respectively. These findings have implications for serodiagnosis and serosurveillance to further our understanding of the epidemiology, transmission and immunopathogenesis in regions where multiple flaviviruses co-circulate.

In agreement with previous reports of cross-reactivities of flavivirus E proteins [24-27], anti-E antibodies were found to cross-react to all six flaviviruses (DENV1, 2 and 4, WNV, ZIKV and YFV) tested in our control panels including pDENV, sDENV, pZIKV, DENV+ZIKV, WNV and YF-17D panels, except that two NHPs and five participants receiving YF-17D vaccine recognized YFV E protein only probably due to generally weak antibody response to live-attenuated vaccine or sampling time ≤2 months or >3−5 years after vaccination (Table S2).

Interestingly, anti-YFV prM antibodies were found in the majority (21/23) of YFV samples tested but only in few from other control panels with an overall sensitivity/specificity of 91.3%/92.5% (Tables 1 and 2). Consistent with our previous report, anti-prM antibodies can discriminate DENV, ZIKV and WNV infections (36). Notably, anti-prM antibodies to one to three DENV serotype were detected in 22/25 of the DENV+ZIKV panel but in 2/23 of the pZIKV panel (*P*<0.0001, two tailed Fisher exact test, Table 1), suggesting that anti-DENV prM antibodies can distinguish these two panels. Although the sensitivities of anti-prM antibodies, ranging from 88,9% (WNV) to 91.3%−91.7% (DENV, ZIKV and YFV) were moderate, the specificities were high (92.5%-99.2%), which was most interesting given that low specificity has been a concern for many serological tests for flaviviruses.

The overall sensitivity/specificity of anti-NS1 antibodies for DENV, ZIKV, WNV, and YFV infections/vaccination were 93.8/51.2%, 100/86.4%, 50.0/99.4%, and 95.7/70.8%, respectively, suggesting that anti-NS1 antibody is not a specific marker for the four flavivirus serocomplexes tested in our assay. Compared with ELISA, our previous study reported that detection of anti-NS1 antibodies in Western blot analysis was less specific, probably due to the presence of cross-reactive anti-NS1 antibodies that recognized linear epitopes in detergent-treated NS1 monomers in Western blot analysis but not NS1 hexamers in solution such as in ELISA [36]. Within the DENV serocomplex, we found that the sDENV panel had higher rates of detecting anti-prM and anti-NS1 antibodies compared with the pDENV panel (98.0% vs. 81.0% and 98.0% vs. 76.2%, respectively, *P*=0.02 and *P*=0.008, Fisher exact test, Table 1). Nonetheless, the difference was insufficient to distinguish these two panels [24,36]. Of note, protein bands corresponding DENV or YFV NS1 protein were recognized by some DENV-naive samples, however, none of the six flavivirus E proteins tested was recognized by these samples (Table 1), suggesting non-specific binding to proteins present in these cell lysates.

The ZIKV outbreak in the Americas has drawn renewed interest in the epidemiology and transmission of ZIKV in other parts of the world (15). Several lines of evidence including documented Zika cases among travelers from Southeast Asia, retrospective analysis of archived samples, and enhanced surveillance suggested that ZIKV has been circulating at a low but sustained level in several countries in Southeast Asia including the Philippines [46-52]. Consistent with these reports, we found approximately 10% of the participants from the Philippines had previous ZIKV or ZIKV and DENV infections [51,52]. The DENV detection rate of 70% among the participants (aged 2−56 years) was generally in agreement with the seroprevalence rate estimated previously [53]. When testing with 48 samples of suspected Zika cases collected between November 2015 and June 2016 in Salvador, we found that all participants (100%) had previous DENV infection and 52.1% (25/48) had previous DENV and ZIKV infections, which was generally in agreement with previous reports of ZIKV seroprevalence during the early phase of ZIKV outbreak in the Northeastern Brazil [54,55]. It is worth noting that the recent YFV outbreak in the Southeastern Brazil started in November 2016, when deaths of NHPs due to YFV infection was reported, followed by human cases since December 2016 with a total of 2237 cases as of June 2019 [10,56,57]; thus, detection of anti-YFV prM antibodies in our participants was unlikely due to the recent YFV outbreak. Our findings that 60.4% (29/48) of participants (aged 15−63 years) had anti-YFV prM antibodies was consistent with the estimated coverage rate of YF vaccination in Brazil (30−70% for individuals aged 15−70 years) and suggested they had previous YF-17D vaccination [58]. Nonetheless, the possibility of exposure during previous YFV outbreaks (2000-2001 and 2008-2009) in the Southeastern and Northeastern states of Brazil cannot be completed ruled out [10,56,57].

Although different neutralization tests including PRNTs, focus reduction neutralization, microneutralization test, reporter viruses, and flow-based neutralization tests, have been developed for flaviviruses, the requirement of labor-intensive work, trained staff, equipment and appropriate biocontainment has limited their applications to reference or research laboratories. Compared with PRNTs, our Western blot analysis is faster (18 h for six viral antigens vs. 5-6 days for PRNTs for each virus), and requires less sample volume (5 µL vs. 128 µL for PRNTs for six antigens or viruses). Moreover, the half-membrane employed in our Western blot analysis can be prepared in advance, stored in a -20°C freezer, and hybridized to serum/plasma up to four months with comparable results (Figure S1), supporting potential application in regional laboratories. It can also be prepared in strips for use as a simple, inexpensive and readily applicable diagnostic test, as exemplified by the first-generation HIV immunoassay [59].

One major challenge of PRNT is that it cannot discriminate multiple flavivirus infections experienced in the past, thus restricting its application for serosurveillance in flavivirus-endemic regions. Despite a previous study reporting different cross-neutralization patterns observed in various ZIKV and DENV infections, whether a defined neutralizing antibody titer (PRNT_50_ titer) can discriminate sDENV and DENV+ZIKV panels or it can be applied to regions where other flaviviruses (YFV, WNV or JEV) are prevalent remains unclear [60]. A recent study in Indonesian Archipelago, a DENV hyperendemic region, revealed multitypic neutralizing antibodies to two or more DENV serotypes and suggested possible ZIKV circulation based on high stringent PRNT_90_ titers, underscoring the difficulty of using PRNT to delineate DENV and ZIKV infections in hyperendemic regions [61]. Other studies used two-step neutralization tests (initial screening by PRNT to ZIKV followed by PRNT to DENV1-4 and ZIKV) [48,49] or combination of IgG ELISA and neutralization test (for positive or equivocal samples) to investigate ZIKV or DENV seroprevalence in endemic regions [62-64]. However, the interpretation of multitypic neutralizing antibody profile remains a challenge.

Our assay using a half membrane with six flavivirus antigens to detect IgG in Western blot analysis can be combined with IgG ELISA to verify those positive or equivocal samples and provide detailed information of infections/exposure of four flavivirus serocomplexes in the past. Given the high specificities of anti-prM antibodies (92.5%-99.2%) in our assay, combination of our assay with other serological tests is unlikely to reduce the overall specificity. Our assay has several potential applications, such as determining flavivirus immune background of participants in a vaccine trial or a seroepidemiological study in endemic regions, and confirming infections with the four flavivirus serocomplexes during surveillance. Our assay can also be employed in retrospective studies of pregnant women with CZS or normal babies to investigate the influence of different ZIKV, DENV, YFV, and/or WNV immune status on pregnancy outcomes. These together would improve our understanding of the epidemiology, immunopathogenesis and complications of ZIKV and DENV in flavivirus-endemic regions.

There were several limitations of this study. First, the sample size in each panel of well-documented flavivirus infections or YF-17D vaccination was relatively small; future studies with larger sample size are warranted to validate these observations. Second, although samples were collected from four months to 31 years after pDENV or sDENV infection and from two months to five years after YF-17D vaccination, samples with longer duration following other flavivirus infections such as ZIKV, DENV+ZIKV and WNV are needed to verify these observations. Third, anti-YFV prM antibodies cannot distinguish YFV natural infection and vaccination with YF-17D, a live-attenuated vaccine. Similarly, anti-DENV prM antibodies cannot discriminate DENV natural infection and vaccinees who received live-attenuated DENV vaccines. Given the availability of other flavivirus vaccines including JEV and tick-borne encephalitis virus vaccines and several ongoing vaccines trials in endemic regions, serological tests that can distinguish flavivirus natural infection and vaccination remain to be exploited in future studies [65,66].

## Data Availability

All data produced in the present work are contained in the manuscript.

## Acknowledgments

We thank Drs. E. Harris at the University of California Berkeley and A. Balmaseda at the Ministry of Health, Managua, Nicaragua for providing samples from Nicaragua, S. L. Stramer at the American Red Cross at Gaithersburg, Maryland for providing samples from blood donors, GJ. Chang at the Center for Disease Control and Prevention at Fort Collins for providing the plasmid expressing YF-17D prM/E proteins, Colorado, and S. Verma at the John A. Burns School of Medicine, University of Hawaii at Manoa for providing WNV-infected cell lysates.

## Author contributions

GHC, YCD and WKW contributed to study design. GHC, YCD and SZH conducted the experiments. GHC, YCD and WKW performed the data analysis. GHC, YCD and WKW had access to underlying data. JJT, AKS, MJ, CP, CB, EMN, PJK, DRDS, DLV, SH, YSH and WKW contributed to reagent or sample collection and funding acquisition. GHC, YCD and WKW contributed to manuscript writing. All authors contributed to the article and approved the submitted version.

## Supplemental Materials

Table S1 and S2

Figure S1

## References

1. Pierson TC, Diamond MS. 2013. Flaviviruses. Knipe DM, Howley PM, eds. Fields virology, 6th ed, Philadelphia: Lippincott William & Wilkins. pp 747-794.

2. Guzman MG, Harris E. Dengue. Lancet. 2015;385:453–465.

3. Bhatt S, Gething PW, Brady OJ, et al. The global distribution and burden of dengue. Nature. 2013;496:504–507.

4. World Health Organization. Dengue and severe dengue. https://www.who.int/news-room/fact-sheets/detail/dengue-and-severe-dengue

5. Halstead SB, Dans LF. Dengue infection and advances in dengue vaccines for children. Lancet Child Adolesc Health. 2019;3:734–741.

6. WHO. Dengue vaccine: WHO position paper - September 2018. Weekly Epidemiological Record. 2018;93:457–476.

7. Sridhar S, Luedtke A, Langevin E, et al. Effect of dengue serostatus on dengue vaccine safety and efficacy. N Engl J Med. 2018;379:327–340.

8. Wilder-Smith A, Smith PG, Luo R, et al. Pre-vaccination screening strategies for the use of the CYD-TDV dengue vaccine: A meeting report. Vaccine. 2019;37:5137–5146.

9. Chen LH, Wilson ME. Yellow fever control: current epidemiology and vaccination strategies. Trop Dis Travel Med Vaccines. 2020;6:1.

10. de Oliveira Figueiredo P, Stoffella-Dutra AG, Barbosa Costa G, et al. Re-emergence of yellow fever in Brazil during 2016-2019: challenges, lessons learned, and perspectives. Viruses. 2020;12:1233.

11. Girard M, Nelson CB, Picot V, et al. Arboviruses: A global public health threat. Vaccine. 2020;38:3989–3994.

12. Jani C, Kakoullis L, Abdallah N, et al. West Nile virus: another emerging arboviral risk for travelers? Curr Infect Dis Rep. 2022;24:117–128.

13. Lessler J, Chaisson LH, Kucirka LM, et al. Assessing the global threat from Zika virus. Science. 2016;353:aaf8160.

14. PAHO, Regional Zika epidemiological update (Americas) - 25 August 2017 [accessed Dec. 1, 2020]. available from: http://www.paho.org/hq/index.php?option=com_content&view=article&id=11599&Itemid=41691&lang=en

15. Musso D, Ko AI, Baud D. Zika virus infection - after the pandemic. N Engl J Med. 2019;381:1444–1457.

16. Anderson KB, Gibbons RV, Thomas SJ, et al. Preexisting Japanese encephalitis virus neutralizing antibodies and increased symptomatic dengue illness in a school-based cohort in Thailand. PLoS Negl Trop Dis. 2011;5:e1311.

17. Rodriguez-Barraquer I, Costa F, Nascimento EJM, et al. Impact of preexisting dengue immunity on Zika virus emergence in a dengue endemic region. Science. 2019;363:607–610.

18. Gordon A, Gresh L, Ojeda S, et al. Prior dengue virus infection and risk of Zika: A pediatric cohort in Nicaragua. PLoS Med. 2019;16:e1002726.

19. Katzelnick LC, Narvaez C, Arguello S, et al. Zika virus infection enhances future risk of severe dengue disease. Science. 2020;369:1123–8.

20. Tsai WY, Lin HE, Wang WK. Complexity of human antibody response to dengue virus: implication for vaccine development. Front Microbiol. 2017;8:1372.

21. Martin DA, Muth DA, Brown T, et al. 2000. Standardization of immunoglobulin M capture enzyme-linked immunosorbent assays for routine diagnosis of arboviral infections. J Clin Microbiol. 2000;38:1823–1826.

22. Johnson AJ, Martin DA, Karabatsos N, et al. Detection of anti-arboviral immunoglobulin G by using a monoclonal antibody-based capture enzyme-linked immunosorbent assay. J Clin Microbiol. 2000;38:1827–1831.

23. Guidance for U.S. Laboratories testing for Zika virus infection. From CDC’s website: http://www.cdc.gov/zika/laboratories/lab-guidance.html.

24. Lai CY, Tsai WY, Lin SR, et al. Antibodies to envelope glycoprotein of dengue virus during the natural course of infection are predominantly cross-reactive and recognize epitopes containing highly conserved residues at the fusion loop of domain II. J Virol. 2008;82:6631–6643.

25. Lanciotti RS, Kosoy OL, Laven JJ, et al. Genetic and serologic properties of Zika virus associated with an epidemic, Yap State, Micronesia, 2007. Emerg Infect Dis. 2008;14:1232-1239.

26. Johnson BW, Kosoy O, Martin DA, et al. West Nile virus infection and serologic response among persons previously vaccinated against yellow fever and Japanese encephalitis viruses. Vector Borne Zoonotic Dis. 2005;5:137–145.

27. Felix AC, Souza NCS, Figueiredo WM, et al. Cross reactivity of commercial anti-dengue immunoassays in patients with acute Zika virus infection. J Med Virol. 2017;89:1477–1479.

28. Steinhagen K, Probst C, Radzimski C, et al. Serodiagnosis of Zika virus (ZIKV) infections by a novel NS1-based ELISA devoid of cross-reactivity with dengue virus antibodies: a multicohort study of assay performance, 2015 to 2016. Euro Surveill. 2016;21:30426.

29. Safronetz D, Sloan A, Stein DR, et al. Evaluation of 5 commercially available Zika virus immunoassays. Emerg Infect Dis. 2017;23:1577–1580.

30. Balmaseda A, Zambrana JV, Collado D, et al. Comparison of four serological methods and two reverse transcription-PCR assays for diagnosis and surveillance of Zika virus infection. J Clin Microbiol. 2018;56:e01785–17.

31. Tsai WY, Youn HH, Brites C, et al. Distinguishing secondary dengue virus infection from Zika virus infection with previous dengue by combination of three simple serological tests. Clin Infect Dis. 2017;65:1829–1836.

32. Tyson J, Tsai WY, Tsai JJ, et al. A high-throughput and multiplex microsphere immunoassay based on non-structural protein 1 can discriminate three flavivirus infections. PLoS Negl Trop Dis. 2019; 13:e0007649.

33. Innis BL. 1997. Antibody responses to dengue virus infection. In Gubler DJ, Kuno G eds: Dengue and dengue hemorrhagic fever. Cambridge, MA: CAB International, pp. 221–244.

34. Halstead SB. Neutralization and antibody-dependent enhancement of dengue viruses. Adv. Virus Res. 2003;60:421–467.

35. Alvarez M, Rodriguez-Roche R, Bernardo L, et al. Dengue hemorrhagic Fever caused by sequential dengue 1–3 virus infections over a long time interval: Havana epidemic, 2001-2002. Am J Trop Med Hyg. 2006;75:1113-1117.

36. Hsieh SC, Tsai WY, Tsai JJ, et al. Identification of anti-premembrane antibody as a serocomplex-specific marker to discriminate Zika, dengue, and West Nile virus infections. J Virol. 2021;95:e0061921.

37. Kuan G, Gordon A, Avilés W, et al. The Nicaraguan pediatric dengue cohort study: study design, methods, use of information technology, and extension to other infectious diseases. Am J Epidemiol. 2009;170:120–129.

38. NarvaezF, Gutierrez G, Perez MA, et al. Evaluation of the Traditional and revised WHO classifications of dengue disease severity. PLoS Negl Trop Dis. 2011;5:e1397.

39. Herrera BB, Tsai WY, Brites C, et al. T cell responses to nonstructural protein 3 distinguish infections by dengue and Zika viruses. mBio. 2018;9: e00755–18.

40. Tyson J, Tsai WY, Tsai JJ, et al. Combination of non-structural protein 1-based enzyme-linked immunosorbent assays can detect and distinguish various dengue virus and Zika virus infections. J Clin Microbiol. 2019;57:e01464–18.

41. Tsai JJ, Liu CK, Tsai WY, et al. Seroprevalence of dengue in two districts of Kaohsiung city after the largest dengue outbreak in Taiwan since world war II. PLoS Negl Trop Dis. 2018;12: e0006879

42. Tsai WY, Chen HL, Tsai JJ, et al. Potent neutralizing human monoclonal antibodies preferentially target mature dengue virus particles: implication for novel strategy of dengue vaccine. J Virol. 2018;92:e00556–18.

43. Dai YC, Sy AK, Jiz M, et al. Identification of prior dengue-naïve Dengvaxia recipients with an increased risk for symptomatic dengue during fever surveillance in the Philippines. Front Immunol 2023;14:1202055.

44. Tsai WY, Driesse K, Tsai JJ, et al. Enzyme-linked immunosorbent assays using virus-like particles containing mutations of conserved residues on envelop protein can distinguish three flavivirus infections. Emerg Microbe Infect. 2020;9:1722–1732.

45. Frey A, Di Canzio J, Zurakowski D. A statistically defined endpoint titer determination method for immunoassays. J. Immunol. Methods. 1998;221:35–41.

46. Duong V, Dussart P, Buchy P. Zika virus in Asia. Int J Infect Dis. 2017;54:121–128.

47. Sasmono RT, Dhenni R, Yohan B, et al. Zika virus seropositivity in 1-4-year-old children, Indonesia, 2014. Emerg Infect Dis. 2018;24:1740-1743.

48. Sasmono RT, Johar E, Yohan B, et al. Spatiotemporal heterogeneity of Zika virus transmission in Indonesia: serosurveillance data from a pediatric population. Am J Trop Med Hyg. 2021;104:2220–2223.

49. Pastorino B, Sengvilaipaseuth O, Chanthongthip A, et al. Low Zika virus seroprevalence in Vientiane, Laos, 2003-2015. Am J Trop Med Hyg. 2019;100:639-642.

50. Ruchusatsawat K, Wongjaroen P, Posanacharoen A, et al. Long-term circulation of Zika virus in Thailand: an observational study. Lancet Infect Dis. 2019;19:439–446.

51. Biggs JR, Sy AK, Brady OJ, et al. Serological evidence of widespread Zika transmission across the Philippines. Viruses. 2021;13:1441.

52. Lonogan K. de Guzman A, Delos Reyes VC, et al. The enhanced Zika surveillance in the Philippines, November 14, 2016–February 28, 2017. Int J Infect Dis. 2020, 101;232–233.

53. L’Azou M, Moureau A, Sarti E, et al. Symptomatic Dengue in Children in 10 Asian and Latin American Countries. N Engl J Med. 2016;374:1155–1166.

54. Netto EM, Moreira-Soto A, Pedroso C, et al. High Zika virus seroprevalence in Salvador, Northeastern Brazil limits the potential for further outbreaks. MBio. 2017;8 e01390–17.

55. Alves LV, Leal CA, Alves JGB. Zika virus seroprevalence in women who gave birth during Zika virus outbreak in Brazil - a prospective observational study. Heliyon. 2020;6:e04817.

56. Rezende IM, Sacchetto L, Munhoz de Mello É, et al. Persistence of yellow fever virus outside the Amazon Basin, causing epidemics in Southeast Brazil, from 2016 to 2018.PLoS Negl Trop Dis. 2018;12:e0006538.

57. Dexheimer Paploski IA, Souza RL, Tauro LB, et al. Epizootic outbreak of yellow fever virus and risk for human disease in Salvador, Brazil. Ann Intern Med. 2018;168:301–302.

58. Shearer FM, Moyes CL, Pigott DM, et al. Global yellow fever vaccination coverage from 1970 to 2016: an adjusted retrospective analysis. Lancet Infect Dis. 2017;17:1209–17.

59. Centers for Disease Control and Prevention and Association of Public Health Laboratories. Laboratory Testing for the Diagnosis of HIV Infection: Updated Recommendations. 2014. http://stacks.cdc.gov/view/cdc/23447.

60. Montoya M, Collins M, Dejnirattisai W, et al. Longitudinal analysis of antibody cross-neutralization following Zika virus and dengue virus infection in Asia and the Americas. J Infect Dis. 2018;218:536–545.

61. Harapan H, Panta K, Michie A, et al. Hyperendemic dengue and possible Zika circulation in the westernmost region of the Indonesian Archipelago. Viruses. 2022;14:219.

62. Nurtop E, Villarroel PMS, Pastorino B, et al. Combination of ELISA screening and seroneutralisation tests to expedite Zika virus seroprevalence studies. Virol J. 2018;15:192.

63. Saba Villarroel PM, Nurtop E, Pastorino B, et al. Zika virus epidemiology in Bolivia: A seroprevalence study in volunteer blood donors. PLoS Negl Trop Dis. 2018;12:e0006239.

64. Lopez AL, Adams C, Ylade M, et al. Determining dengue virus serostatus by indirect IgG ELISA compared with focus reduction neutralisation test in children in Cebu, Philippines: a prospective population-based study. Lancet Glob Health. 2021;9:e44-e51.

65. Simmons G, Stone M, Busch MP. 2018. Arbovirus diagnostics: from bad to worse due to expanding dengue virus vaccination and Zika virus epidemics. Clin Infect Dis. 2018; 66:1181–13.

66. Munoz-Jordan JL. Diagnosis of Zika virus infections: challenges and opportunities. J Infect Dis. 2017;216:S951–S956.

