## Supplemental Figure 1 and Tables 1 and 2 for "Detection of anti-premembrane antibody as a specific marker of four flavivirus serocomplexes and its application to serosurveillance in endemic regions"

# FS1

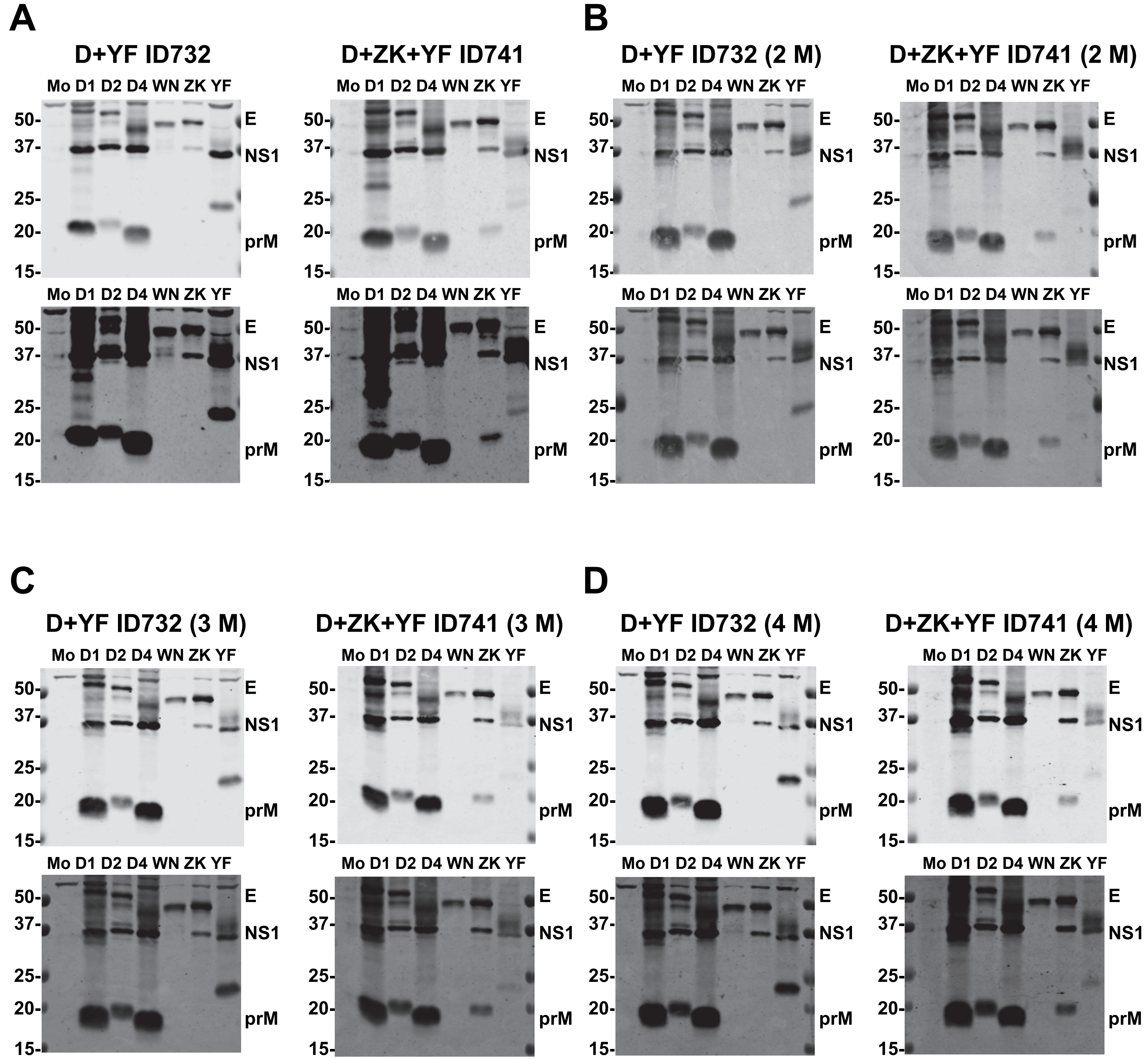


**Figure S1.** Results of Western blot analysis using pre-prepared and stored half-membrane. Serum samples ID732 with previous DENV and ZIKV infections (D+ZK) and ID741 with previous DENV, ZIKV and YFV infections/vaccination (D+ZK+YF) were hybridized with half-membranes immediately after transfer (A), or with those stored in -20°C freezer for 2 months (B), 3 months (C) and 4 months (D). Both short (upper) and long (lower) exposures of each gel are shown. The positions of E, NS1 and prM protein bands are indicated. The size of molecular weight markers is shown in kDa. Mo: mock, D1: DENV1, D2: DENV2, D4: DENV4, WN: WNV, ZK: ZIKV, and YF: YF-17D.

**Table S1.** Numbers, sources and basic information of different serum/plasma panels with confirmed flavivirus infections or vaccination

| Panel[a] | No. of samples | Sampling time[b] | Sources (No. of samples), Country and year [reference] | Confirmation methods |
|---|---|---|---|---|
| pDENV | 21 | 6M−31Y | seroprevalence study (17), Taiwan, 2015−16 [41] | Microeutralization test[c] |
| | | unknown | ZIKV study (4), Brazil, 2016−17 [39] | Microeutralization test[c] |
| sDENV | 50 | 4M−29Y | seroprevalence study (29), Taiwan, 2015−16 [41] | Microeutralization test[c] |
| | | unknown | ZIKV study (21), Brazil, 2016−17 [39] | Microeutralization test[c] |
| pZIKV | 23 | 6−8M | ZIKV study (18), Nicaragua, 2016−17 [40] | RT-PCR |
| | | 5M−11M | ZIKV study (5), Brazil, 2016−17 [39] | Microeutralization test[c] |
| DENV+ZIKV | 25 | 6−8M | ZIKV study (13), Nicaragua, 2016−17 [40] | RT-PCR |
| | | 5M−11M | ZIKV study (12), Brazil, 2016−17 [39] | Microeutralization test[c] |
| YF-17D | 23 | <1M−5Y | YF-17D study (10), U.S. 1990−2016 [43] | vaccination history |
| | | unknown | ZIKV study (9), Brazil, 2016−17 [39] | vaccination history |
| | | 1M−1.5Y | NHP study (4), BEI Resources, 2011−12 | vaccination history |
| WNV | 18 | <1M | WNV study (18), U.S. 2006−15 [40] | TMA[d] |
| DENV-naive | 29 | NA | seroprevalence study (29), Taiwan, 2015−16 [41] | Microeutralization test[c] or multiple ELISAs |

[a]pDENV, primary DENV infection; sDENV, secondary DENV infection; WNV, WNV infection; pZIKV, primary ZIKV infection; DENV+ZIKV, previous DENV and ZIKV infections; YF-17D, YF-17D vaccination; NA, not applicable.
[b]Sampling time post-symptom onset, YF-17D vaccination, or TMA test.
[c]Microneutralization test as described previously [39,42].
[d]Index samples tested positive for WNV transcription-mediated amplification (TMA), IgM and IgG from blood donors at the American Red Cross [40].

**Table S2.** Viral proteins recognized by different panels in Western blot analysis

| Panel[a]/ID | NS1[b] | | | | | | | prM[b] | | | | | | | E[c] |
|---|---|---|---|---|---|---|---|---|---|---|---|---|---|---|---|
| pDENV[d] | D1 | D2 | D4 | any D | WNV | ZIKV | YFV | D1 | D2 | D4 | any D | WNV | ZIKV | YFV | D1,2,4,W,Z,Y[c] |
| S037.20Y | 1 | | | 1 | | | | 1 | | 1 | 1 | | | | 1 |
| S049.30Y | 1 | 1 | | 1 | | | | | | | | | | | 1 |
| S213.31Y | 1 | | 1 | 1 | | | 1 | 1 | | 1 | 1 | | | | 1 |
| S281.16M | 1 | 1 | 1 | 1 | | | | 1 | 1 | 1 | 1 | | | | 1 |
| S287.27Y | 1 | | 1 | 1 | | | 1 | 1 | | 1 | 1 | | | | 1 |
| S298.6M | 1 | 1 | 1 | 1 | | | | 1 | | 1 | 1 | | | | 1 |
| S016 | | | | | | | | | | | | | | | 1 |
| S017 | 1 | 1 | 1 | 1 | | | | 1 | | 1 | 1 | | | | 1 |
| S019 | | | | | | | | 1 | | 1 | 1 | | | | 1 |
| S047 | | | | | | | | | | | | | | | 1 |
| S065 | 1 | | 1 | 1 | | | | 1 | | 1 | 1 | | | | 1 |
| S020 | 1 | | | 1 | | | | 1 | | | 1 | | | | 1 |
| S025 | 1 | 1 | | 1 | | | | 1 | | | 1 | | | | 1 |
| S087 | | | | | | | | | | | | | | 1 | 1 |
| S386.1Y | 1 | 1 | | 1 | | | 1 | 1 | | 1 | 1 | | | | 1 |
| S433.29Y | 1 | 1 | 1 | 1 | | | 1 | 1 | 1 | 1 | 1 | | | | 1 |
| S425.3Y | 1 | 1 | | 1 | | | 1 | 1 | | 1 | 1 | | | | 1 |
| ZK0966 | 1 | 1 | 1 | 1 | | | NA | 1 | 1 | 1 | 1 | | | NA | 1 |
| ZK0980 | | | | | | | NA | 1 | 1 | 1 | 1 | | | NA | 1 |
| ZK0995 | 1 | | | 1 | | | NA | 1 | 1 | 1 | 1 | | | NA | 1 |
| ZK0997 | 1 | 1 | 1 | 1 | | | NA | 1 | 1 | 1 | 1 | | | NA | 1 |
| Total n=21 | 16 | 10 | 9 | 16 | 0 | 0 | 5 | 17 | 6 | 15 | 17 | 0 | 0 | 1 | 21 |
| sDENV[d] | D1 | D2 | D4 | any D | WNV | ZIKV | YFV | D1 | D2 | D4 | any D | WNV | ZIKV | YFV | D1,2,4,W,Z,Y[c] |
| S006 | | | 1 | 1 | | | | 1 | | | 1 | | | | 1 |
| S008 | 1 | 1 | 1 | 1 | | 1 | | 1 | 1 | 1 | 1 | | | | 1 |
| N229.23Y | 1 | | | 1 | | | | 1 | | | 1 | | | | 1 |
| N258.17M | 1 | 1 | | 1 | | | | 1 | | | 1 | | | | 1 |

| | | | | | | | | | | | | | |
|---|---|---|---|---|---|---|---|---|---|---|---|---|---|
| S051.25Y | 1 | 1 | | 1 | | | 1 | | 1 | 1 | | | 1 |
| S217.16M | 1 | | | 1 | | | 1 | | 1 | 1 | | | 1 |
| S075 | 1 | 1 | | 1 | | 1 | 1 | 1 | 1 | 1 | 1 | | 1 |
| S080 | 1 | | | 1 | | | 1 | | | 1 | | | 1 |
| S289 | 1 | 1 | | 1 | | | 1 | 1 | 1 | 1 | | | 1 |
| S034 | 1 | | | 1 | | | | | | | | | 1 |
| S035 | 1 | | | 1 | | 1 | 1 | | 1 | 1 | | | 1 |
| S038 | 1 | 1 | 1 | 1 | | 1 | 1 | 1 | 1 | 1 | | 1 | 1 |
| S059 | | 1 | | 1 | | | 1 | 1 | 1 | 1 | | | 1 |
| S069 | | | | | | | 1 | | 1 | 1 | | | 1 |
| S264 | 1 | 1 | 1 | 1 | | | 1 | | 1 | 1 | | | 1 |
| S028.4M | 1 | 1 | | 1 | | | 1 | | | 1 | | | 1 |
| S044.1Y | 1 | | 0 | 1 | | | 1 | 1 | 1 | 1 | | | 1 |
| S053 | 1 | 1 | | 1 | 1 | | 1 | | | 1 | | | 1 |
| S055 | 1 | 0 | | 1 | | | 1 | | 1 | 1 | | | 1 |
| S066.1Y | 1 | 1 | 1 | 1 | | | 1 | 1 | 1 | 1 | | | 1 |
| S089.1Y | 1 | 1 | 1 | 1 | | | 1 | 1 | 1 | 1 | | | 1 |
| N040.25Y | 1 | | | 1 | | | 1 | | 1 | 1 | | | 1 |
| N438.15Y | 1 | 1 | | 1 | | | 1 | | | 1 | | 1 | 1 |
| N406.29Y | 1 | 1 | | 1 | | 1 | 1 | | | 1 | | | 1 |
| S370.28Y | 1 | 1 | | 1 | | | 1 | 1 | 1 | 1 | | | 1 |
| S444.26Y | 1 | 1 | | 1 | | | 1 | | 1 | 1 | | | 1 |
| S473.2Y | 1 | 0 | | 1 | | | 1 | 1 | 1 | 1 | | | 1 |
| NH003.24Y | 1 | | | 1 | | 1 | 1 | | 1 | 1 | | | 1 |
| NH005.29Y | 1 | | | 1 | | 1 | 1 | | 1 | 1 | | | 1 |
| ZK0976 | 1 | 1 | 1 | 1 | 1 | NA | 1 | 1 | 1 | 1 | | NA | 1 |
| ZK0986 | 1 | 1 | 1 | 1 | 1 | NA | 1 | 1 | 1 | 1 | | NA | 1 |
| ZK0967 | 1 | 1 | 1 | 1 | 1 | NA | 1 | 1 | 1 | 1 | | NA | 1 |
| ZK0969 | 1 | 1 | 1 | 1 | 1 | NA | 1 | 1 | 1 | 1 | | NA | 1 |
| ZK0971 | 1 | 1 | 1 | 1 | 1 | NA | 1 | 1 | 1 | 1 | | NA | 1 |

| | | | | | | | | | | | | | | | |
|---|---|---|---|---|---|---|---|---|---|---|---|---|---|---|---|
| ZK0977 | 1 | 1 | 1 | 1 | | 1 | NA | 1 | 1 | 1 | 1 | | | NA | 1 |
| ZK0983 | 1 | 1 | 1 | 1 | | | NA | 1 | 1 | 1 | 1 | | | NA | 1 |
| ZL0985 | 1 | 1 | 1 | 1 | | | NA | 1 | 1 | 1 | 1 | | | NA | 1 |
| ZK0988 | 1 | 1 | 1 | 1 | | 1 | NA | 1 | 1 | 1 | 1 | | | NA | 1 |
| ZK0992 | 1 | 1 | 1 | 1 | | 1 | NA | 1 | 1 | 1 | 1 | | | NA | 1 |
| ZK0994 | 1 | 1 | 1 | 1 | | 1 | NA | 1 | 1 | 1 | 1 | | | NA | 1 |
| ZK1001 | 1 | 1 | 1 | 1 | | 1 | NA | 1 | 1 | 1 | 1 | | | NA | 1 |
| ZK1010 | 1 | 1 | 1 | 1 | | 1 | NA | 1 | 1 | 1 | 1 | | | NA | 1 |
| ZK1013 | 1 | 1 | 1 | 1 | | | NA | 1 | 1 | 1 | 1 | | | NA | 1 |
| ZK0973 | 1 | 1 | 1 | 1 | | | NA | 1 | 1 | 1 | 1 | | | NA | 1 |
| ZK0974 | 1 | 1 | | 1 | | | NA | 1 | 1 | 1 | 1 | | | NA | 1 |
| ZK1002 | 1 | 1 | | 1 | | | NA | 1 | 1 | 1 | 1 | | | NA | 1 |
| ZK1003 | 1 | 1 | 1 | 1 | | | NA | 1 | 1 | 1 | 1 | | | NA | 1 |
| ZK1007 | 1 | 1 | 1 | 1 | | | NA | 1 | 1 | 1 | 1 | | | NA | 1 |
| ZK1016 | 1 | 1 | 1 | 1 | | | NA | 1 | 1 | 1 | 1 | | | NA | 1 |
| ZK0990 | 1 | 1 | 1 | 1 | | | NA | 1 | 1 | 1 | 1 | | | NA | 1 |
| Total n=50 | 47 | 37 | 25 | 49 | 0 | 13 | 6 | 49 | 31 | 41 | 49 | 0 | 1 | 2 | 50 |
| pZIKV[d] | D1 | D2 | D4 | any D | WNV | ZIKV | YFV | D1 | D2 | D4 | any D | WNV | ZIKV | YFV | D1,2,4,W,Z,Y[c] |
| 7613.7M | | | 1 | 1 | | 1 | | | | | | | 1 | | 1 |
| 5839.7M | 1 | 1 | 1 | 1 | | 1 | | | | | | | 1 | | 1 |
| 5966.7M | 1 | 1 | 1 | 1 | | 1 | 1 | | | | | | 1 | | 1 |
| 7434.8M | | | | | | 1 | | | | | | | 1 | | 1 |
| 7658.8M | 1 | 1 | 1 | 1 | | 1 | | | | | | | 1 | | 1 |
| 5248.7M | 1 | 1 | 1 | 1 | | 1 | | | | | | | 1 | | 1 |
| 4041.7M | | | | | | 1 | | | | | | | 1 | | 1 |
| 5893.8M | 1 | 1 | 1 | 1 | | 1 | | | | | | | 1 | | 1 |
| 4445.7M | 1 | 1 | 1 | 1 | | 1 | | | | 1 | | | 1 | | 1 |
| 7304.7M | 1 | 1 | 1 | 1 | | 1 | | 1 | | 1 | | | 1 | | 1 |
| 5953.7M | 1 | 1 | 1 | 1 | | 1 | | | | | | | 1 | | 1 |
| 6763.7M | 1 | 1 | 1 | 1 | | 1 | | | | | | | 1 | 1 | 1 |

| | | | | | | | | | | | | | | |
|---|---|---|---|---|---|---|---|---|---|---|---|---|---|---|
| 6563.6M | 1 | 1 | 1 | 1 | | 1 | | | | | | | 1 | | 1 |
| 6669.6M | 1 | 1 | 1 | 1 | | 1 | | | | | | | 1 | | 1 |
| 7253.6M | 1 | 1 | 1 | 1 | | 1 | | | | | | | 1 | | 1 |
| 6697.6M | 1 | 1 | 1 | 1 | | 1 | 1 | | | | | | 1 | 1 | 1 |
| 7444.7M | 1 | 1 | 1 | 1 | | 1 | | | | | | | 1 | | 1 |
| 6248.7M | 1 | 1 | 1 | 1 | | 1 | 1 | | | | | | 1 | | 1 |
| ZK0979.5-11M | 1 | 1 | 1 | 1 | | 1 | NA | | | | | | 1 | NA | 1 |
| ZK0993.5-11M | | | | | | 1 | NA | | | | | | 1 | NA | 1 |
| ZK0998.5-11M | 1 | 1 | | 1 | | 1 | NA | | | | | | 1 | NA | 1 |
| ZL1006.5-11M | 1 | 1 | | 1 | | 1 | NA | | | | | | 1 | NA | 1 |
| ZK0996.5-11M | | | 1 | 1 | | 1 | NA | | | | | | 1 | NA | 1 |
| Total n=23 | 18 | 18 | 18 | 20 | 0 | 23 | 3 | 1 | 0 | 2 | 2 | 0 | 23 | 2 | 23 |
| DENV+ZIKV[d] | D1 | D2 | D4 | any D | WNV | ZIKV | YFV | D1 | D2 | D4 | any D | WNV | ZIKV | YFV | D1,2,4,W,Z,Y[c] |
| 7487.7M | 1 | 1 | 1 | 1 | | 1 | 1 | 1 | | 1 | 1 | | 1 | | 1 |
| 6775.7M | 1 | 1 | 1 | 1 | | 1 | | 1 | | 1 | 1 | | 1 | | 1 |
| 4749.7M | 1 | 1 | 1 | 1 | | 1 | 1 | 1 | | 1 | 1 | 1 | 1 | | 1 |
| 7933.7M | 1 | 1 | | 1 | | 1 | | 1 | | | 1 | | 1 | | 1 |
| 5550.8M | 1 | 1 | 1 | 1 | | 1 | 1 | | | | | | 1 | 1 | 1 |
| 7208.8M | 1 | 1 | 1 | 1 | | 1 | 1 | 1 | 1 | 1 | 1 | | 1 | | 1 |
| 6364.7M | 1 | 1 | 1 | 1 | | 1 | 1 | 1 | 1 | 1 | 1 | | 1 | | 1 |
| 4069.7M | 1 | 1 | 1 | 1 | | 1 | 1 | 1 | 1 | 1 | 1 | | 1 | | 1 |
| 5962.7M | 1 | 1 | 1 | 1 | 1 | 1 | | | | | | 1 | 1 | | 1 |
| 5355.7M | 1 | 1 | 1 | 1 | | 1 | 1 | | | | | 1 | 1 | | 1 |
| 2049.6M | 1 | 1 | 1 | 1 | | 1 | 1 | 1 | 1 | 1 | 1 | | 1 | | 1 |
| 2056.6M | 1 | 1 | 1 | 1 | | 1 | 1 | 1 | | 1 | 1 | | 1 | 1 | 1 |
| 2076.6M | 1 | 1 | 1 | 1 | | 1 | 1 | 1 | 1 | 1 | 1 | | 1 | | 1 |
| ZK0972.5-11M | 1 | 1 | 1 | 1 | | 1 | NA | 1 | 1 | 1 | 1 | | 1 | NA | 1 |
| ZK0975.5-11M | 1 | 1 | 1 | 1 | | 1 | NA | 1 | 1 | 1 | 1 | | 1 | NA | 1 |
| ZK0989.5-11M | 1 | 1 | 1 | 1 | | 1 | NA | 1 | 1 | 1 | 1 | | 1 | NA | 1 |
| ZK0991.5-11M | 1 | 1 | 1 | 1 | | 1 | NA | 1 | 1 | 1 | 1 | | | NA | 1 |

| | | | | | | | | | | | | | | |
|---|---|---|---|---|---|---|---|---|---|---|---|---|---|---|
| ZK1000.5-11M | 1 | 1 | 1 | 1 | | 1 | NA | 1 | 1 | 1 | 1 | | | NA | 1 |
| ZK1009.5-11M | 1 | 1 | 1 | 1 | | 1 | NA | 1 | 1 | 1 | 1 | | 1 | NA | 1 |
| ZK1011.5-11M | 1 | 1 | 1 | 1 | | 1 | NA | 1 | 1 | 1 | 1 | | | NA | 1 |
| ZK1012.5-11M | 1 | 1 | 1 | 1 | | 1 | NA | 1 | 1 | 1 | 1 | | 1 | NA | 1 |
| ZK1014.5-11M | 1 | 1 | 1 | 1 | | 1 | NA | 1 | 1 | 1 | 1 | | 1 | NA | 1 |
| ZK1015.5-11M | 1 | 1 | 1 | 1 | | 1 | NA | 1 | 1 | 1 | 1 | | | NA | 1 |
| ZK0968.5-11M | 1 | 1 | 1 | 1 | | 1 | NA | 1 | 1 | 1 | 1 | | 1 | NA | 1 |
| ZK0984.5-11M | 1 | 1 | 1 | 1 | | 1 | NA | 1 | 1 | 1 | 1 | | 1 | NA | 1 |
| Total n=25 | 25 | 25 | 24 | 25 | 1 | 25 | 10 | 22 | 17 | 21 | 22 | 3 | 21 | 2 | 25 |
| YF-17D[d] | D1 | D2 | D4 | any D | WNV | ZIKV | YFV | D1 | D2 | D4 | any D | WNV | ZIKV | YFV | D1,2,4,W,Z,Y[c] |
| YFV0 | | | | | | | 1 | | | | | | | 1 | 1 |
| YFV1.3M | | | | | | | 1 | | | | | | | 1 | 1 |
| YFV5.5Y | | | | | | | 1 | | | | | | | 1 | Y |
| YFV14.5Y | | | | | | | 1 | | | | | | | 1 | Y |
| YFV17.2M | | | | | | | 1 | | | | | | | 1 | Y |
| YFV18.15M | | | | | | | 1 | | | | | | | 1 | 1 |
| YFV19.4M | | | | | | | 1 | | | | | | | 1 | 1 |
| YFV20.4M | 1 | 1 | 1 | 1 | | | 1 | 1 | 1 | 1 | 1 | | | 1 | 1 |
| YFV12.3Y | | | | | | | 1 | | | | | | | 1 | Y |
| YFV2<1M | | | | | | | 1 | | | | | | | 1 | Y |
| NHP#12.1-18M | | | | | | | 1 | | | | | | | 1 | 1 |
| NHP#29.1-18M | | | | | | | 1 | | | | | | | 1 | Y,W,Z |
| NHP#30.1-18M | | | | | | | 1 | | | | | | | 1 | Y |
| NHP#41.1-18M | | | | | | | 1 | | | | | | | 1 | 1 |
| ZK745 | NA | NA | NA | NA | | NA | 1 | NA | NA | NA | NA | | NA | 1 | 1 |
| ZK750 | NA | NA | NA | NA | | NA | 1 | NA | NA | NA | NA | | NA | 1 | 1 |
| ZK757 | NA | NA | NA | NA | | NA | 1 | NA | NA | NA | NA | | NA | 1 | 1 |
| ZK765 | NA | NA | NA | NA | | NA | 1 | NA | NA | NA | NA | | NA | 1 | 1 |
| ZK772 | NA | NA | NA | NA | | NA | | NA | NA | NA | NA | | NA | | 1 |
| ZK779 | NA | NA | NA | NA | | NA | 1 | NA | NA | NA | NA | | NA | | 1 |

| | | | | | | | | | | | | | | |
|---|---|---|---|---|---|---|---|---|---|---|---|---|---|---|
| ZK066 | NA | NA | NA | NA | | NA | 1 | NA | NA | NA | NA | | NA | 1 | 1 |
| ZK343 | NA | NA | NA | NA | | NA | 1 | NA | NA | NA | NA | | NA | 1 | 1 |
| ZK083 | NA | NA | NA | NA | | NA | 1 | NA | NA | NA | NA | | NA | 1 | 1 |
| Total n=23 | 1 | 1 | 1 | 1 | 0 | 0 | 22 | 1 | 1 | 1 | 1 | 0 | 0 | 21 | 23 |
| WNV[e] | D1 | D2 | D4 | any D | WNV | ZIKV | YFV | D1 | D2 | D4 | any D | WNV | ZIKV | YFV | D1,2,4,W,Z,Y[c] |
| WR 3052<1M | | | | | | | NA | | | | | 1 | | NA | 1 |
| WR 2994<1M | | | 1 | 1 | | | NA | | | | | 1 | | NA | 1 |
| WR 2931<1M | | | | | | | NA | | | | | | | NA | 1 |
| WR 2787<1M | 1 | 1 | 1 | 1 | | | NA | | | | | 1 | | NA | 1 |
| WR 2726<1M | | | | | | | NA | | | | | 1 | | NA | 1 |
| WR 2728<1M | | | 1 | 1 | 1 | | NA | | | | | 1 | | NA | 1 |
| WR 2659<1M | | | 1 | 1 | 1 | | NA | | | | | 1 | | NA | 1 |
| WR 2573<1M | 1 | 1 | 1 | 1 | 1 | 1 | NA | | | | | 1 | | NA | 1 |
| WR 2441<1M | 1 | 1 | 1 | 1 | | | NA | | | | | 1 | | NA | 1 |
| WR 2419<1M | | 1 | 1 | 1 | 1 | 1 | NA | | | | | 1 | | NA | 1 |
| WR 2096<1M | | 1 | 1 | 1 | 1 | 1 | NA | | | | | 1 | | NA | 1 |
| WR 1595<1M | 1 | 1 | 1 | 1 | 1 | | NA | | | | | 1 | | NA | 1 |
| WR 1569<1M | | | | | | | NA | | | | | | | NA | 1 |
| WR 1561<1M | 1 | 1 | 1 | 1 | 1 | 1 | NA | | | | | 1 | | NA | 1 |
| WR 1518<1M | 1 | 1 | 1 | 1 | | | NA | | | | | 1 | | NA | 1 |
| WR 1478<1M | 1 | 1 | 1 | 1 | | | NA | | | | | 1 | | NA | 1 |
| WR 1428<1M | 1 | 1 | 1 | 1 | 1 | 1 | NA | | | | | 1 | | NA | 1 |
| WR 1346<1M | 1 | 1 | 1 | 1 | 1 | | NA | | | | | 1 | | NA | 1 |
| Total n=18 | 9 | 11 | 14 | 14 | 9 | 5 | 0 | 0 | 0 | 0 | 0 | 16 | 0 | 0 | 18 |
| DENV-naive | D1 | D2 | D4 | any D | WNV | ZIKV | YFV | D1 | D2 | D4 | any D | WNV | ZIKV | YFV | D1,2,4,W,Z,Y[c] |
| N002 | | | | | | | 1 | | | | | | | | |
| N003 | | | | | | | | | | | | | | | |
| N004 | | | 1 | 1 | | | 1 | | | | | | | | |
| N005 | | | | | | | | | | | | | | | |
| N036 | | | | | | | | | | | | | | | |

| | | | | | | | | | | | | | | | |
|---|---|---|---|---|---|---|---|---|---|---|---|---|---|---|---|
| N021 | | | | | | | | | | | | | | | |
| N029 | | | | | | | | | | | | | | | |
| N037 | | | | | | | 1 | | | | | | | | |
| N043 | | | | | | | 1 | | | | | | | | |
| N044 | | | | | | | | | | | | | | | |
| N047 | | | | | | | | | | | | | | | |
| N048 | 1 | | 1 | 1 | | | | | | | | | | | |
| N050 | 1 | | 1 | 1 | | | | | | | | | | | |
| N051 | 1 | | 1 | 1 | | | 1 | | | | | | | | |
| N053 | | | | | | | | | | | | | | | |
| N055 | 1 | | | 1 | | | | | | | | | | | |
| N045 | | | | | | | | | | | | | | | |
| N054 | | | | | | | | | | | | | | | |
| N056 | | | | | | | | | | | | | | | |
| N060 | | | | | | | | | | | | | | | |
| Neg P1 | | | | | | | | | | | | | | | |
| Neg P2 | | | | | | | | | | | | | | | |
| Neg P3 | | | | | | | | | | | | | | | |
| N006 | | | | | | | 1 | | | | | | | | |
| N008 | | | 1 | 1 | | | 1 | | | | | | | 1 | |
| N009 | | | | | | | | | | | | | | | |
| N012 | | | | | | | | | | | | | | | |
| N016 | | | | | | | | | | | | | | | |
| Neg P4 | | | | | | | | | | | | | | | |
| Total n=29 | 4 | 0 | 5 | 6 | 0 | 0 | 7 | 0 | 0 | 0 | 0 | 0 | 0 | 1 | 0 |

[a]pDENV, primary DENV infection; sDENV, secondary DENV infection; pZIKV, primary ZIKV infection; DENV+ZIKV, previous DENV and ZIKV infections; WNV, WNV infection; YF-17D, YF-17D vaccination
[b]NS1, nonstructural protein 1; prM, premembrane; 1, positive to the indicated NS1 or prM protein band.
[c]E, envelope; D1,2,4, DENV1,2, 4; W, WNV; Z, ZIKV; Y, YFV; 1, positive to all six E protein bands tested; Y or other letters, positive to YFV or other E proteins, respectively.
[d]Known sampling time post-symptom onset, YF-17D vaccination, or TMA test was shown after sample ID.
[e]Index samples tested positive for WNV transcription-mediated amplification (TMA), IgM and IgG from blood donors at the American Red Cross [40].
